# Metagenomic Sequencing for Wastewater-Based Surveillance: Modeling and Experimental Approaches for Determining Limit of Detection

**DOI:** 10.64898/2026.08.18.26360688

**Authors:** Amy Xiao, Katherine Besse, Daisy Connors, Trina Vian, Jim Stylinski, Anthony Mannion, Joe Lacirignola

**Affiliations:** Lincoln Laboratory, Massachusetts Institute of Technology, Lexington, MA, USA

## Abstract

Since the COVID-19 pandemic, wastewater-based surveillance (WBS) has emerged as a key approach to assess community-level health and the evolution of pathogens. To date, most established WBS systems focus on polymerase chain reaction (PCR) based detection and targeted sequencing of known pathogens because these approaches are well-accepted and include amplification of pathogen target sequences of interest thereby enabling lower limits of detection. Metagenomic next-generation sequencing (mNGS) is a promising approach to enable pathogen detection and surveillance beyond predefined pathogen lists, but its regular application to WBS has not been yet widely adopted because many key performance characteristics are not well-understood, including limit of detection (LOD) and false positive/negative rates. This paper describes a computational analysis to estimate the operational LOD of various sequencing approaches using a simplified model of a local wastewater (WW) system involving a military base. This paper also presents findings from two types of experiments: 1) laboratory-spiked, those for which *Atlantibacter subterraneus* (Asub) is introduced into real-world WW samples in a laboratory setting, and 2) system-spiked, those for which Asub is introduced at a source location of a real-world WW system. Findings indicate that mNGS detection performance varies with sequencing method and the data analysis process. In addition, findings indicate that site-specific method characterization should be used when implementing mNGS for WBS because sites can have different WW system configurations, background organisms and sequencing inhibitors.

## INTRODUCTION

Widely implemented during the COVID-19 pandemic, wastewater-based surveillance (WBS) enables community-level disease monitoring and provides an early warning of pathogen spread and evolution. Many groups have shown that SARS-CoV-2 detections in wastewater preceded widespread clinical reporting. ^1,2,3,4,5^ It has been previously demonstrated that SARS-CoV-2 viral copy numbers in wastewater were higher than expected from confirmed clinical cases,^6,7,8,9,10^ suggesting that WBS may form a more comprehensive picture of community health and could provide early warning of outbreaks, due to the contribution of asymptomatic shedders or those who do not seek clinical care. Currently, public health agencies, including the United States Centers for Disease Control and Prevention (CDC), use quantitative polymerase chain reaction (qPCR) and targeted sequencing to monitor for the presence of a pre-determined list of pathogens.^11^ Both qPCR-based and targeted sequencing methods enable the tracking of temporal trends in pathogen levels via longitudinal analysis.^12,13^

While targeted methods have been routinely used to provide public health information, the COVID-19 pandemic demonstrates the challenges that a rapidly evolving pathogen poses for PCR-based detection methods. Furthermore, there is an ever-growing risk of novel and emerging pathogens due to human encroachment of natural habitats, climate change, rapid global travel and increasing use of artificial intelligence and access to biotechnology^14,15,16^ These risk factors underscore the need to detect potential pathogens in a sample without prior pathogen knowledge. For this effort, we focus on metagenomic next generation sequencing (mNGS) (or ‘shotgun’ sequencing) as the technical approach to broadly characterize the nucleic acid present in WW samples for organism identification.

mNGS can been used to characterize the microbial composition of a sample, enabling extensive human and environmental microbiome studies.^17,18,19,20,21,22,23,24^ However, the operational use of mNGS (i.e., serving as a basis for public health decision making, development of infectious disease diagnostics, and environmental biological pathogen detection^25,26,27^) has been hindered by several challenges, including complex non-standardized workflows, lack of understanding of detection metrics such as the limit of detection (LOD), difficulty relating pathogen sequencing reads to the associated disease risks and the overall cost per sample analysis. As a result, there are currently no FDA-approved mNGS diagnostic tests,^28^ and mNGS approaches have not been widely adopted for operational wastewater surveillance.

Several recent journal articles have provided insights into LOD and other mNGS performance metrics.^29, 30,31,32,33,^ The studies typically vary in terms of the workflow, sequencing type and depth, what is used to generate a gold standard result, and how detection metrics are calculated. As a result, it is difficult to compare results across different efforts. To understand the potential advantage of mNGS-based wastewater surveillance approaches, both modeling and experimental approaches are necessary to assess feasibility and actionability. To this end, Grimm et al. used statistical modelling approaches to quantify the sensitivity and cost of mNGS-based wastewater surveillance for viral pathogen detection, finding that relative abundance of SARS-CoV-2 varied by orders of magnitude across studies and that hybridization panels can greatly reduce cost and improve sensitivity.^34^ A pilot study on the implementation of wastewater surveillance at skilled-nursing facilities conducted dye-based tracer studies to assess feasibility of additional sites.^35^ Sonnenwald et al. conducted fluorescent dye tracing at the sewer network level and developed a simplified hydraulic modeling approach to predict solute concentrations across the network following an injection of dye.^36^ Kohle et al. flushed 10^13^ CFU of *Staphylococcus hyicus* in a sewershed of ∼36,000 people and showed a sensitivity of 16-140 sick people based on shotgun metagenomic sequencing from samples collected at the wastewater treatment plant.^37^ These studies emphasize the importance of characterizing performance metrics like sensitivity and limit of detection to demonstrate the utility of shotgun metagenomic sequencing for WBS-informed public health decisions.

In this study, we explore methods to characterize the operational LOD for wastewater mNGS and various sampling architectures using both theoretical and experimentally-derived results. We define the operational LOD as the lowest concentration of an analyte that can be reliably distinguished from a blank sample in the operationally-relevant context (here, a wastewater system). To assess the theoretical performance of mNGS, inform field experiments and derive cost estimates, we built a computational model of a local wastewater system network to estimate the performance of qPCR and mNGS to enable the potential detection of a range of pathogen-infected cases shedding into the wastewater system of a military base. The computational analysis was conducted for a set of individual cases (as well as higher levels of cases) shedding into the wastewater system. This paper presents the performance estimates for the detection of a single case, as this is the most stressing scenario and one that could potentially enable the earliest detection of an emerging novel pathogen. To validate model findings, we conducted a laboratory-based spike test to assess the feasibility of detection with untargeted metagenomic sequencing in wastewater. We then conducted wastewater system-spike experiments to measure the operational mNGS LOD for a local wastewater system for different sequencing technologies and analysis methods. Collectively, the combination of computational and laboratory insights and validation through experimental results provides a method for defining mNGS performance for wastewater systems.

## MATERIALS AND METHODS

### Computational model development and assumptions

To inform LOD organism-spike experiments, an estimate of genomic target concentration and dwell time as a function of sampling location was needed to guide study design. A WW network model was developed and configured to emulate Hanscom Air Force Base (HAFB) where system spike experiments would be performed. HAFB consists of multiple building types including varying types of commercial and residential structures. The complex base WW network was simplified to reduce computational complexity and time. Simplifying assumptions to reduce complexity included neglecting stormwater runoff and infiltration, combining adjacent commercial or residential areas as a single source where practical and defining "system manholes” (i.e., discrete sampling points within the model) at main sewage line junctions. Finer sampling resolution was retained within the MIT Lincoln Laboratory complex to support follow-on experimentation. A schematic of the simplified network is shown in Figure S1. Pipe material type, diameter and approximate run length were identified directly from specifications provided by HAFB personnel.

In the simplified model, a 1D advection and dispersion equation was coupled with series routing logic and exponential decay of target concentration over time. The modeling of advection and dispersion of solutes within a fluid is given by the equation:

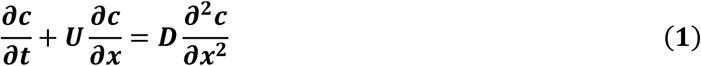

where c = concentration (gene-copies/mL), U = velocity (m/s), D = dispersion coefficient. The concentration with respect to distance along x and time, *t,* is:

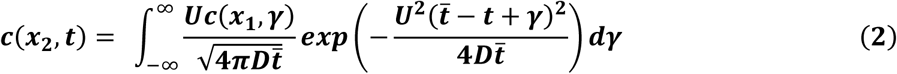

To model the degradation of RNA with time in WW, the SARS-CoV-2 virus was selected as a representative pathogen due to extensive WW characterization that occurred during the COVID-19 pandemic. The concentration of SARS-CoV-2 RNA is modeled using ^38^

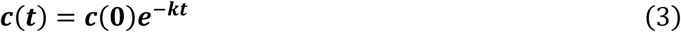

Additional simplifying assumptions (related to solute transport and degradation) include: steady-state gravity flow within pipes, pressure main between wet-well pump stations is treated as time delay, longitudinal dispersion is characterized by the Manning equation ^39^,

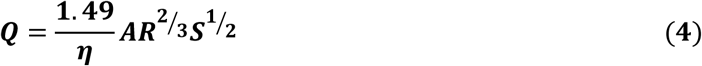

Where

Q = discharge,
*η* = Manning’s roughness coefficient,
A = area,
R= hydraulic radius,
S= slope.

target organism is injected into the system at the beginning of a simulation, lower station pump runs continuously, and sampling performed at any point in the network is lossless. Other model parameters are listed in Table 1. Water usage estimates were obtained from the Massachusetts state environmental code for establishment type^40^ and summarized in Table S1.

**Table 1:** Key parameters used in the Matlab^®^ simulation of the HAFB wastewater network.

| Category | Parameter | Value(s) |
| --- | --- | --- |
| Pathogen | Shed rate (SARS-CoV-2) | $10^{7.53} - 10^{9.27}$ gc/g-feces<sup>41</sup> |
| | WW degradation coefficient | $1.752 \text{ day}^{-1}$<sup>42</sup> |
| Source | Mean fecal mass | $120 \text{ g}$<sup>43</sup> |
|  | Pulse duration | 10 - 40sec |
| | Pulse frequency | $1.2 \text{ day}^{-1}$ |
| Hydraulic | Manning roughness | $0.01 - 0.019 \text{ s/m}^{1/3}$ |
|  | Pipe slope | 0.02 m/m |
| | Dispersion coefficient | $0.009 - 0.374 \text{ m}^2/\text{s}$ |
|  | Pump station wet well volume | 5200 gal (max 10,000 gal) |
| Simulation | Time step | 5 s |
|  | Simulation duration | 5 h |

### Estimated Detection Platform Operational LOD

Concentration profiles were generated by the model in units of gene copies per unit volume (gc/mL) to allow easy conversion between target organisms with differing genome sizes. LODs for various sequencing platforms are not specified by the manufacturer, as LOD varies with factors including sample background, target, lab processing methods and data analysis methods; rather, a minimum input mass is recommended by the manufacturer for sequencing library preparation. To estimate the detection technology’s LOD requires the conversion from minimum input mass to gc/mL. Assumptions about sample volumes at various stages of the preparation process, genome size of the target organism and amount of target contained in the input mass can yield a theoretical minimum input for mNGS. In this study, using SARS-CoV-2 as a representative pathogen, and accounting for pragmatic recovery efficiencies along the way, this input mass is adjusted and referred to as the *realistic minimum input*. These assumptions are listed in Table 2. Table 3 summarizes the estimated theoretical and realistic platform minimum inputs for three different detection technologies.

**Table 2:** Key assumptions for mass to gc/mL conversion.

|  |  |
| --- | --- |
| SARS-CoV-2 genome size<sup>44</sup> | ~29,900 RNA bases |
| Overall recovery efficiency<br>at *each* of 4 steps of processing | 10% |
| Target mass contributing<br>to minimum input mass | 1% |
| Sample volume processed for<br>sequencing | 300 mL |
| Reaction volume for qPCR | 50 µL |
| Amplification bias | Not considered |
| SARS-CoV-2 shed rate<sup>45</sup> | 10<sup>7</sup> - 10<sup>9</sup> gc/g-feces |
| Target mass per flush<br>(120 g stool containing 10<sup>9</sup> gc/g)<sup>46</sup> | 1.2 × 10<sup>11</sup> gc |

**Table 3:** Estimated minimum input in gc/mL for select detection technologies.

| Technology | Library Prep Protocol | Minimum Input | Theoretical Estimated Minimum Input (target gc/mL) | Realistic Estimated Minimum Input (target gc/mL) |
| --- | --- | --- | --- | --- |
| Oxford Nanopore MinION (sequencing) | Ligation Sequencing Kit | 1,000 ng<sup>47</sup> | $2.1 \times 10^7$ | $2.1 \times 10^{10}$ |
| Illumina NextSeq550 (sequencing) | Illumina DNA Prep | 1 ng<sup>48</sup> | $2.1 \times 10^4$ | $2.1 \times 10^7$ |
| Applied Biosystems QuantStudio7 (quantitative PCR) | N/A | 80 gc/mL<sup>49</sup> | 80 | 80 |

### Preparation of spike in bacteria for laboratory and system spike experiments

*Atlantibacter subterraneus* (Asub - formerly known as *Salmonella subterranea*) was selected for the laboratory and system spike experiments due to its ease of propagation in large volumes and low background presence in Kraken2 analysis of previous HAFB wastewater samples. A freeze-dried bacterial strain of Asub was purchased from ATCC (ATCC BAA-836) and rehydrated following ATCC handling instructions. The rehydrated bacteria were streaked onto Tryptic Soy Agar (TSA) plates to obtain single colonies. Individual colonies were used to inoculate multiple Tryptic Soy Broth (TSB) cultures, which were incubated overnight at 35 ℃ with aeration. Glycerol stocks were prepared by adding 500 µL of the culture to 500 µL of sterile filtered 50% glycerol in cryovials for storage at -80 ℃.

For preparation of bacteria for system spike in experiments, bacteria growth was scaled up across multiple days in shake flasks and incubated at 35 ℃ shaking from 150-250 RPM based on the size of the flask. On the starting date, multiple 5 mL cultures were started from glycerol stocks of Asub and incubated for 24 hours before pooling and adding to a 150 mL TSB culture. These cultures were incubated for another 24 hours before back dilution and split into multiple 500 mL cultures. After another 24 hours of growth, approximately 2 L of bacteria were available for the system spike-in. The cumulative time to grow up this material was approximately 72 hours. The optical density at 600 nm (OD600) of the culture was measured throughout the growing process. The procedure for preparing bacteria for the laboratory spike was similar to above, but due to lower total culture volumes needed, cultures were incubated for 24 hours only.

### Heat inactivation of bacteria before system spikes

Bacteria were required to be 100% inactivated before spiking into the sewer system based on our organization’s Environment, Health, and Safety (EHS) department and public works guidelines. Heat treatment at 90 ℃ for 15 minutes in a water bath was shown to fully inactivate bacteria and minimally impact downstream molecular detection. The heat-inactivated bacteria were streaked onto TSA plates and left to incubate overnight at 35 ℃ to verify inactivation.

### Quantification of bacteria preparations

A qPCR assay was designed to quantify genomic copy numbers of Asub. Primer and probe sequences are listed in Table S2. This assay was tested against a positive control DNA fragment (DedA - Integrated DNA Technologies) and nucleic acids extracted from culture using the Qiagen DNeasy PowerSoil Kit (Qiagen 12855-100). To create a standard curve, twelve samples from a 10-fold dilution series of the extracted culture in nuclease free water were tested in duplicate using the Luna Universal qPCR Master Mix (NEB M3003X). 2 µL of sample was added to 10 µL of master mix, 1 µL of the PrimeTime qPCR custom assay (20×) and 7 µL of nuclease free water. Cycling parameters were as follows: 95 ℃ for 20 s, followed by 40 cycles of denaturing at 95 ℃ for 3 s and annealing/extension at 60 ℃ for 30 s on a Thermo Fisher QuantStudio 7 Pro. To determine the LOD of the qPCR assay, 20 replicates were tested and the concentration at which 19/20 replicates were positive was identified as the assay LOD (13 copies per reaction, Cq = 37.26)

The same 10-fold dilution series of the extracted culture was quantified by digital PCR (Qiagen QIAcuity) using the assays outlined above, the QIAcuity OneStep Advanced Probe Kit (Qiagen 250131) and the QIAcuity Nanoplate 8.5k 96-well plates (Qiagen 250021). 2 µL of sample were added to 3 µL One Step Advanced Probe Master Mix (4×), 0.6 µL of PrimeTime qPCR custom assay (20×), 1.5 µL of GC Enhancer and 4.9 µL of nuclease free water. Cycling parameters were as follows: 95 ℃ for 3 min, followed by 40 cycles of denaturing at 95 ℃ for 5 s and annealing/extension at 60 ℃ for 30 s. Standard curves were created by combining the results from the dPCR and qPCR experiments. The original concentration of the *Asub* culture was estimated by using the standard plate count method. A serial dilution of the culture was created and 100 µL of each dilution were plated on TSA plates and left to incubate overnight at 35 ℃. Colonies were enumerated and the colony-forming units (CFU)/mL in the starting culture was calculated. 1 mL of the culture was extracted using the Qiagen DNeasy PowerSoil Kit (Qiagen 12855-100), resulting in a final elution volume of 50 µL. With 2 µl added to each qPCR reaction, the CFU represented in each qPCR reaction was calculated and plotted against the qPCR cycle threshold values.

### Laboratory spikes of bacteria into de-ionized water (DI) water and wastewater backgrounds

A 250 mL volume of wastewater was collected from a wastewater pump station in Lexington, MA. Samples were heat-inactivated within 15 min of collection at 60 °C for 60 min in a water bath. Asub was cultured and quantified as described above and spiked into 40 mL aliquots of either DI water or wastewater in a 10× dilution series. Nucleic acid extraction was conducted immediately using the Promega Wizard® Enviro Total Nucleic Acid Kit (Promega, A2991), with 40 mL input per the manufacturer’s instructions. A 40 μL volume was used to elute the nucleic acids. All elutions were run through the *OneStep* PCR Inhibitor Removal Kit (Zymo Research D6031). Post-extraction elutions were quantified using the Qubit 1X dsDNA assay (ThermoFisher Scientific Q33231). qPCR assays targeting Asub and Tomato brown rugose fruit virus *(ToBRFV)* were conducted, and samples were sent for Illumina DNA sequencing by SeqCenter (Pittsburgh, PA, USA).

### System spikes of bacteria into local wastewater network

Wastewater system spike-ins were conducted after acquiring approvals from the EHS team as well as local and greater Boston public works departments. Preliminary dye tests were performed to track timing of material from release point to collection point. 500 mL of fluorescent dye (USABlueBook® Sewer Tracing Dye, Yellow-Green Liquid) was released from the designated point and tracked at the collection point for transit time and visibility. The dye was determined not to interfere with downstream molecular analyses of the wastewater samples. For bacterial spikes, 500 mL of dye was mixed with 500-1250 mL of quantified heat-inactivated bacteria and released at the designated point, followed by 10 flushes. Wastewater samples were collected at the pump station after approximately 30 min; one 250 mL sample every minute until the dye was no longer visible. Eight bacterial spikes were conducted on four different days. Wastewater samples were transported to the lab, and sample processing and nucleic acid extraction were conducted as described above.

### Illumina library preparation and sequencing

Extracted wastewater nucleic acids were sent to SeqCenter (Pittsburg, PA, USA) for Illumina library preparation and sequencing using the Illumina DNA Prep kit and custom IDT 10bp unique dual indices (UDI) with a target insert size of 280 base pairs (bp). Illumina sequencing was performed on an Illumina NovaSeq X Plus sequencer, producing 2×151 bp paired-end reads. Demultiplexing, quality control, and adapter trimming were performed with bcl-convert1 (v4.2.4).

### ONT library preparation and sequencing

Libraries were prepared for long-read sequencing using the ONT Native Barcoding Kit 24 V14 (SQK-NBD114.24) kit, per manufacturer’s instructions. Samples were normalized to the lowest concentration and pooled after the addition of barcodes. Libraries were quantified using the Qubit assay (dsDNA HS, ThermoFisher) per the manufacturer’s instructions. Pooled ONT libraries were sequenced using a dedicated PromethION flow cell at SeqCenter (Pittsburgh, PA, USA).

### Bioinformatic analysis

Adaptor sequences and low-quality bases in Illumina reads were trimmed using BBduk^50^ under default parameters (ktrim=r k=23 mink=11 hdist=1 tpe tbo). ONT pod5 files were basecalled and demultiplexed with dorado v1.1.0 and the dna_r10.4.1_e8.2_400bps_hac@v4.3.0 model. ONT reads were filtered with NanoFilt^51^ to an average Phred score of 10 per read and minimum read length of 100 bp. Average data output post-QC for Illumina samples was 53 Gbp per sample (SD: 9.7 Gbp) and average data output post-QC for ONT samples was 3.4 Gbp per sample (SD: 1.4 Gbp). To facilitate comparisons between samples with varying sequencing output, each sample was subsampled to the lowest output found in the dataset (∼1.2 Gbp per sample). A custom Kraken2^52^ database composed of GTDB bacterial species, viral genomes from NCBI, CheckV, and EsViritu databases and eukaryotic pathogens from EuPathDB was used for read-based taxonomic classification. Minimap2^53^ was used for read mapping to the Asub reference genome (https://genomes.atcc.org/genomes/c52b51a4132d47f0) or a combined reference that included the Asub genome and the *Atlantibacter hermanii* genome (NCBI RefSeq GCF_016027855) to understand the effect of including near-neighbors. The “sr” and “map-ont” parameters were used to map Illumina and ONT reads, respectively. Samtools^54^ was used to calculate coverage and depth with mapping quality (min-MQ = 30) and base quality (min-BQ = 30) thresholds. Total mapped bases were calculated from the samtools depth output. GOTTCHA2^55^ was used as another method to conduct species-specific read mapping. Bioinformatic analyses were conducted using high-performance computing resources from the MIT Lincoln Laboratory Supercomputing Center. ^56^

### Calculating the Operational Limit of Detection (LOD) for mNGS-based WBS

Bases mapped to the target by each bioinformatic tool were normalized by the number of bases post-QC and multiplied by 1,000,000 to obtain mapped bases per million (MBPM). Within each spike experiment, the MBPM of spiked samples was divided by the MBPM of the no-spike background samples to obtain MBPM over background, which is analogous to a signal to noise ratio (SNR). A threshold was established by computing the average MBPM over background of non-spiked samples ± 3 standard deviations to account for the high variability of wastewater samples. The operational LOD was determined to be the concentration of spiked target where MBPM over background exceeded the threshold consistently across all 4 spike experiments. In cases where there was no target detection in the blank, mapped bases = 1 was used for the analysis.

#### Permutation Testing for MBPM over background of Illumina vs ONT sequencing

For each bioinformatic analysis method, samples with paired Illumina and ONT sequencing were selected for permutation testing. The test statistic for the observed data was calculated as follows:

1. Calculation of the difference between the log of the Illumina MBPM over background and the log of the ONT MBPM over background. Log transform prior to differencing was chosen to account for the fact that MBPM over background is a ratio.
2. Average of the difference within spike experiments, to account for the grouped structure
3. Average of the averages across spike experiments, to account for the grouped structure Furthermore, 20,000 permutations of the sign of the differences were conducted and the test statistic was calculated as described above to form a null distribution. The two-tailed p-value was calculated as the number of permuted statistics that were equal to or more extreme than the observed test statistic. When no permutations resulted in a test statistic more extreme than the observed test statistic, the p-value is reported as p<5.0 × 10^-5^ (1/20,000).

## RESULTS

### WW model-based sampling architecture assessment suggests that qPCR monitoring is more sensitive than mNGS for base-level sampling

Three different sampling approaches were examined: monitoring a large installation while sampling at key aggregation points only (two samples each from lower and upper pump station wet well), sampling multiple branches within the installation to improve source localization (ten samples) and sampling outside of each building (100+ samples) to provide the highest level of spatial resolution. Several model simulations were performed varying the number of infected contributors and location sources. 71 scenarios examined a single infected person at each of the simplified model nodes. Estimated detection results for the single shedder scenarios under the three different sampling approaches are shown in Figure 1.A qPCR detection platform shows an estimated minimum input of ∼10^2^ gc/mL, while estimated values of the Illumina and ONT MinION platforms show ∼10^7^ and 10^11^ respectively. With qPCR as a detection platform, the base can be effectively monitored from two sampling locations with the exception being contributions from sources at the most distal ends of the system. Randomly sampling at branches may result in detection with some sequencing platforms although full network coverage is lost unless every branch is included. Random sampling from each building yields high enough concentrations to be detected robustly.

**Figure 1:**
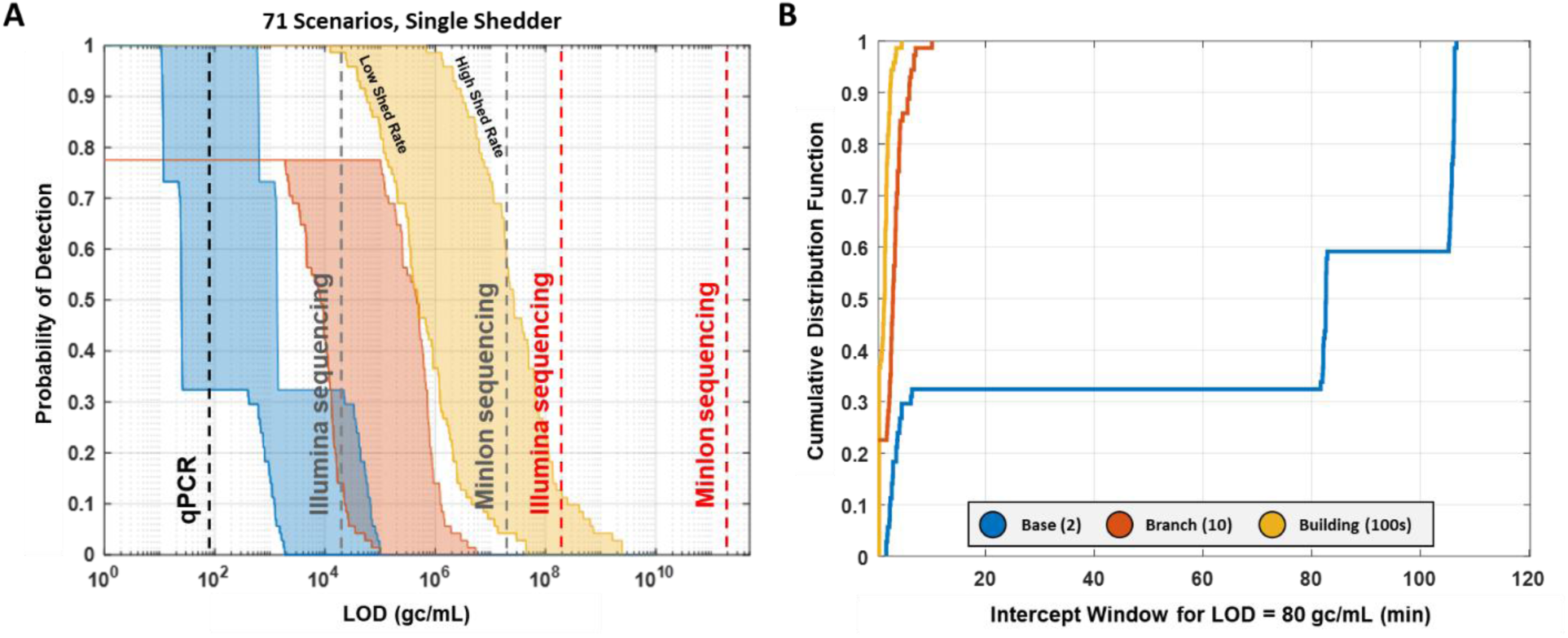
Computational model enables estimation of wastewater surveillance feasibility of different sampling architectures and detection technologies. (A) LOD (estimated mass input) as a function of probability of detection (detectable scenarios) across single shedder simulations. Vertical lines denote estimated detection threshold ranges with a particular detection platform (subject to the assumptions described in the Methods section) at high and low shed rates. Gray = theoretical, red = realistic estimated minimum input. (B) Amount of time in minutes that concentration is above qPCR threshold for the different sampling approaches with a single high shedder at various locations within the network.

Successful detection also requires that the sample can be effectively collected for analysis. The amount of time a sample is above LOD (i.e., dwell time or window to intercept detectable sample) is shown for the most sensitive detection platform (qPCR) in Figure 1. Although overall sample concentrations are lower at the pump station wet wells, they persist much longer than that of the highly transitory pipes and manholes within the system. Unless nearly continuous sampling was implemented, the target could be missed entirely if sampling from pipes and manholes.

Table S3 contains a summary of parameters that affect an overall sampling architecture: sample numbers (a driver of program cost) and detection (ability to acquire sample and detect target). The ONT MinION platform was excluded as no scenario with realistic estimated minimum input was predicted to be detectable. These initial modeling estimates helped to guide when and where to sample within the HAFB wastewater network as well as inform expectations for detection with different mNGS platforms. The model developed can be updated with actual dilution and transit times should a more accurate predictive capability be required. The model can also be adapted quickly to represent other wastewater networks.

### Lab-based spike of A. subterraneus cells into DI water and wastewater sample matrix reveal different LODs for qPCR, Illumina, and ONT sequencing

The computational model highlighted different LODs for qPCR and sequencing but was based on extensive assumptions. To empirically determine the operational LOD, we conducted lab-based spike experiments where whole Asub cells were spiked into wastewater and DI water prior to extraction and molecular detection. The operational LOD of qPCR for DI water and wastewater spikes was determined to be between 150 – 1,500 CFU/mL over background (Figure 2A). This result was consistent with the qPCR assay validation testing, which showed an assay LOD of 13 copies per reaction, corresponding to Cq=37.26. The targeted nature of qPCR suggests that the background matrix should not impact the LOD of the assay. The qPCR detections of lab-based spiked wastewater samples were used to generate a standard curve which could be used to estimate the concentration of the spiked target in the later wastewater system spike experiments (Figure S4).

**Figure 2:**
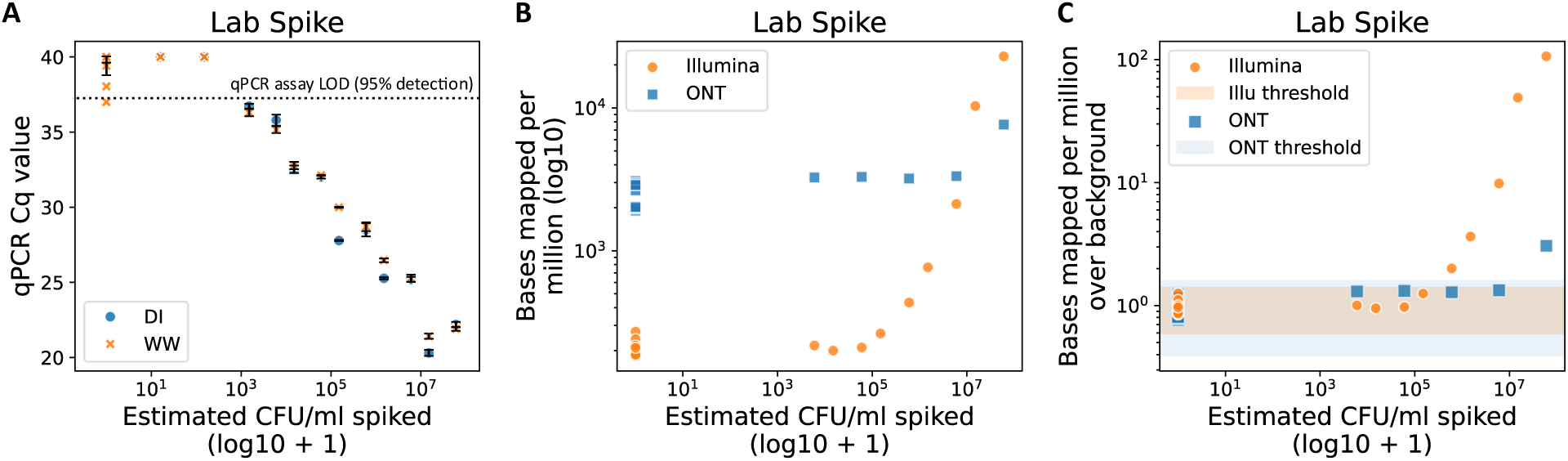
Laboratory spike-in experiments. **(A)** qPCR detection of Asub cells spiked into DI water and wastewater backgrounds. Asub cells were quantified by plating, then spiked into DI water and wastewater samples before nucleic acid extraction and qPCR. qPCR assay validation showed an assay LOD of 13 copies/reaction, corresponding to Cq=37.26. Samples with no qPCR detection were assigned Cq=40. X-axis has a pseudocount of 1 added to visualize blanks on a log scale. **(B)** mNGS detection of Asub cells spiked into DI water and wastewater in the lab on Illumina and ONT platforms. Y-axis represents the number of bases mapped to the Asub reference genome by minimap2 normalized by the total number of bases after quality control, multiplied by a factor of 10^6^. X-axis has a pseudocount of 1 added to visualize blanks on a log scale. **(C)** mNGS detection of *Asub* cells spiked into DI water and wastewater in the lab on Illumina and ONT platforms. Y-axis represents the ratio between the MBPM of the sample compared to the blank. X-axis has a pseudocount of 1 added to visualize blanks on a log scale. Shaded regions in light orange and light blue represent the average MBPM of the blank samples +/- 3 standard deviations for Illumina and ONT, respectively. CFU: colony forming units. DI: deionized water. WW: wastewater. Cq: quantification cycle. MBPM: mapped bases per million, number of bases mapped to the target organism normalized by the number of bases after sequencing read quality control and multiplied by a factor of 10^6^

Wastewater samples spiked with Asub cells were also characterized with mNGS on Illumina and ONT platforms. Illumina data showed a higher target signal than ONT data at high spike concentrations as measured by mapped bases per million (MBPM) (Figure 2B). ONT data showed a higher background signal than Illumina data, where samples without any target spike-in showed ∼3000 MBPM (Figure 2B). Taken together, these observations highlight that Illumina has a higher signal to noise ratio than ONT for this experiment (Figure 2C). The threshold for detection was set as the average MBPM of no-spike wastewater samples + 3 standard deviations. Based on these thresholds, the operational LOD of Illumina sequencing was determined to be between 1.5 × 10^5^ and 6.0 × 10^5^ CFU/mL wastewater, whereas the operational LOD of ONT sequencing was determined to be between 6.0 × 10^5^ and 6.0 × 10^6^ CFU/mL wastewater.

### qPCR detection for wastewater system spike shows background interference

The lab spiking results did not consider the loss of material and dilution that occurs in a real-world sewer network. To understand how the sewer network can influence the operational LOD, we ran several experiments where we spiked the HAFB sewer system with Asub combined with fluorescent dye. The dye and microbial spike were shown to transit the system together (Figure S2). qPCR detection from the system spike experiments is shown in Figure 3. The qPCR assay can detect ∼300 CFU/mL in wastewater, consistent with the lab-based spike results, but endogenous Asub signal in the wastewater background makes the spiked signal indistinguishable from noise until ∼1.3 × 10^5^ – 3.6 × 10^5^ CFU/mL wastewater. This operational LOD is higher than that reported for the lab-based spike (150 – 1,500 CFU/mL wastewater, Figure 2A), which could be due to variable wastewater backgrounds during the months that the system spike experiments were conducted.

**Figure 3:**
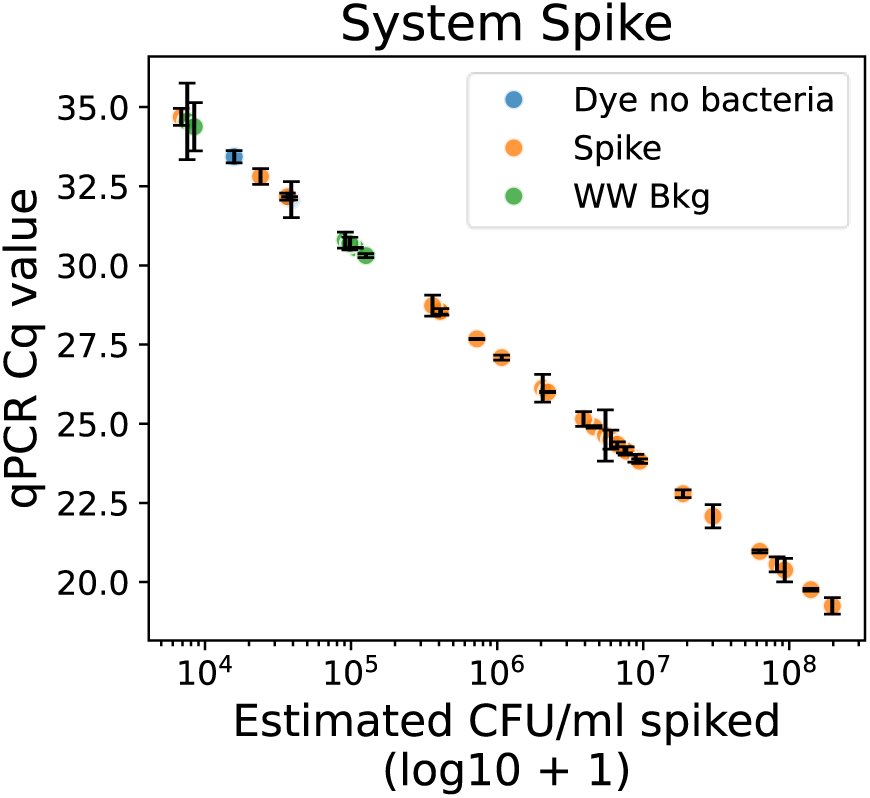
qPCR detection of wastewater system spike. Asub was flushed into the wastewater system. Wastewater samples were collected at the pump station, assayed with qPCR, and CFU/mL in the sample was estimated based on the standard curve in Figure S4. WW bkg: wastewater background before target was spiked. Dye no bacteria: wastewater background with fluorescent dye only, without target bacteria. Spike: samples collected after target bacteria were spiked in the sewer system. CFU: colony forming units. Cq: quantification cycle

### Bioinformatic methods that use species-specific databases enhance signal to noise ratio in mNGS detection of spiked target in wastewater

Because different bioinformatic methods have been shown to have different performance characteristics^57^, we used different bioinformatic methods to analyze the wastewater system spike samples and understand the effect on mNGS detection. We measure detection with the estimated operational LOD as well as the MBPM over background metric, which is analogous to a signal to noise ratio (SNR). Minimap2 mapping of reads to the Asub genome alone showed that detection above background became consistent after the spike concentration exceeded ∼7.6 × 10^6^ CFU/mL wastewater for Illumina data and ∼6.3 × 10^7^ CFU/mL wastewater for ONT data (Figure S5, Figure S6). We further investigated the effect of adding an *A. hermannii* (Aherm) reference genome in addition to the target Asub to the mapping process. The minimap2 results with a two-genome reference showed slightly improved estimated operational LOD for ONT (Figure 4A) and slightly improved SNR compared to the results with only the target genome (Figure S5, Figure S6). As an alternative method, Kraken2 was used to investigate the effect of taxonomic classification with a broad database. With Kraken2, operational LOD decreased to ∼4.1 × 10^5^ CFU/mL wastewater for Illumina data and ∼9.5 × 10^6^ CFU/mL wastewater for ONT data (Figure 4A, Figure S5, Figure S6). Kraken2 further increased SNR for Illumina and ONT data (Figure 4B). Finally, GOTTCHA2 with a broad database was used to map reads against species-specific markers. While operational LOD for GOTTCHA2 analysis was similar to Kraken2 analysis for both Illumina and ONT (Figure 4A), GOTTCHA2 had the highest SNR of all assessed methods (Figure 4B, Figure S5, Figure S6). There is a statistically significant difference in the average MBPM over background for Illumina-sequenced samples compared to ONT-sequenced samples (two-tailed permutation test, Figure 4B). We observe that the MBPM over background metric is higher for Illumina (Figure 4B,D) and the operational LOD is lower for Illumina (Figure 4A,C), with and without subsampling the data to match the lowest ONT sample’s sequencing output.

**Figure 4:**
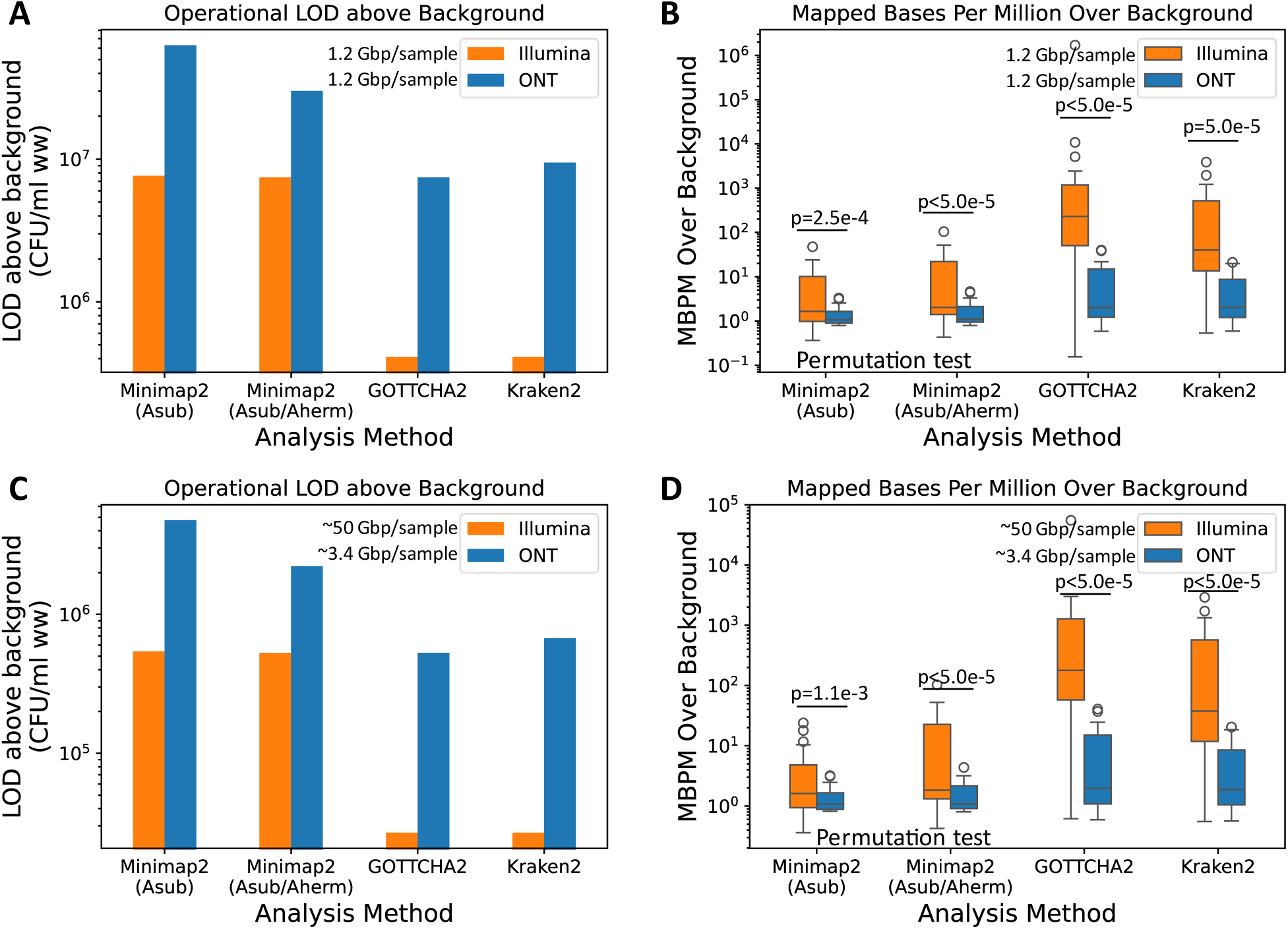
Summary of LOD and SNR for different sequencing and bioinformatic methods, with and without subsampling. **(A)** Limit of detection ranges identified through system-spike experiments for Illumina and ONT datasets analyzed with different bioinformatic methods, all samples subsampled to 1.2 Gbp sequencing output. **(B)** Distribution of MBPM over background values for spike samples, sequenced with Illumina and ONT and analyzed with different bioinformatic methods, all samples subsampled to 1.2 Gbp sequencing output. **(C)** Limit of detection ranges identified through system-spike experiments for Illumina and ONT datasets analyzed with different bioinformatic methods, all samples subsampled to 1.2 Gbp sequencing output, without subsampling. **(D)** Distribution of MBPM over background values for spike samples, sequenced with Illumina and ONT and analyzed with different bioinformatic methods, without subsampling. LOD: limit of detection. SNR: signal to noise ratio. MBPM: mapped bases per million, number of bases mapped to the target organism normalized by the number of bases after sequencing read quality control and multiplied by a factor of 10^6^. Asub: *Atlantibacter subterraneus*. Aherm: *Atlantibacter hermannii*.

Using the lowest dilution factor for a spiked sample (∼5000×, measured by fluorescent dye), the shed rate of Salmonella in animal models (∼10^2^ – 10^7^ CFU/g feces),^58^ and assuming 1 g feces corresponds to 1 mL, the lowest estimated operational LOD (∼4.1 × 10^5^ CFU/mL) corresponds to 2 × 10^2^ – 2 × 10^7^ people with Salmonella infection. This estimate assumes that all material is discharged from one point in the system at once, which is unlikely. Further modeling would be necessary to include variation related to outbreak progression and random toilet usage.

## DISCUSSION

There is a rising interest in using increasingly agnostic detection methods such as mNGS to detect and identify pathogens, including those that may be genetically modified, emerging or novel. However, applying mNGS to operational WBS requires an understanding of its performance metrics at each location and how mNGS information can be used to make actionable decisions. The LOD for a detection system is critical information because it helps inform what a detection means in terms of quantity of potential pathogen in the WW system. This information can then be used to infer how many people may be infected and thus the severity of the pathogen spread. Knowledge of system LOD enables the decision maker to determine the appropriate level of response and risk for a particular community. Here, we describe a process for establishing an mNGS LOD for wastewater surveillance at a military base through computational modeling and experimentally-derived results.

A simplified hydraulic model based on SARS-CoV-2 predicted that mNGS would be less sensitive than qPCR for detecting single cases. The computational model included many simplifications of the variations in system design parameters, flow rates, sampling type, sample processing methodology and sequencing methods. We chose many parameters based on vendor information, available research, and knowledge of the system gained by design documents. Even with all this information, it is difficult to create an exact duplication of the wastewater system due to complex aspects of the hydraulic environment, including the system turbulence, inflow and infiltration, degree of biofilm built up on pipe walls, and variations in daily usage. Potential improvements to the model include estimating inflow and infiltration, adding a time-dependent water usage parameter, and fine-tuning wastewater system design parameters based on dye testing results. The modeling framework described here can be used as a preliminary feasibility assessment when establishing new WBS locations, although location-specific inputs, including wastewater system design and flow rates, will be necessary to tailor results.

Laboratory and wastewater system-based spike experiments assessed how the wastewater sample matrix influences detection limits due to existing microbial content and assay interferents. Operational LOD is primarily affected by the level and variability of target detected in the background sample. Operational LODs for mNGS methods were equivalent to or higher than the qPCR operational LOD, depending upon sequencing and bioinformatics methods. Illumina sequencing outperformed ONT sequencing, with a ∼70× lower operational LOD for some analysis methods. Elevated operational LOD for ONT could be caused by inaccurate read mapping. ONT has been shown to have a higher sequencing error rate than Illumina^59^, which could increase background and decrease target-specific mapping. We observed that the application of the minimap2 algorithm to long vs. short reads generated from identical input samples resulted in a log difference in the signal to noise ratio for some methods, which could be due to different default parameters within the software. Finally, whereas ONT sequencing typically has longer read lengths that could increase confidence in target-specific mapping, nucleic acids could degrade in the wastewater due to environmental and chemical factors as well as during sample processing manipulations. We observe that average ONT read lengths from the spike samples are similar to those of Illumina (Figure S3), which may offset potential advantages of long reads for species-specific identification.

The differences between the model-predicted and the experimentally-derived operational LODs can be attributed to several factors. First, the model assumptions were based on the SARS-CoV-2 virus (genome size ∼29,900 bases), whereas the experiments were conducted with the Gram-negative bacteria, Asub (genome size ∼4.8 Mb). Differences in target organism affect recovery rates during sample processing as well as the relative mass of the target genome compared to the total wastewater nucleic acid output^60^. Accounting for differences in genome length and dsDNA vs ssRNA, the same number of genome copies of Asub is expected to contribute ∼318× more nucleic acid content than SARS-CoV-2. The SARS-CoV-2- based model did not consider how the presence of near-neighbor organisms in the wastewater background would elevate the operational LOD, whereas the experimental results showed non-negligible background signal depending on analysis method. Second, the model assumed that 1% relative abundance of the target among the background was necessary for a confident sequencing detection without specifically considering sequencing output or bioinformatic analysis method. The experimental samples were sequenced deeply and subsampled to 1.2 Gbp per sample for fair comparison to the lowest output sample, and multiple analysis methods were used, many of which require less than 1% relative abundance for a detection.

As mNGS is increasingly incorporated into WBS efforts, it is vital to understand how decision makers would like to use this information to make choices. Discussions with the larger wastewater biosurveillance community (civilian and military) suggest that agnostic detection and identification capabilities for early warning would likely be employed alongside targeted quantitative methods particularly when so much is still unknown about the capability. To move towards operationality, a distributed WBS architecture using mNGS benefits from: standardized protocols (sampling, processing and analysis pipelines), information quality measures (contextual LOD and identification confidence) and knowledge of the normal WW background per region (contributing populations, dilution, diversity trends and inhibitors). Site-specific verification of best methods should be conducted when standing up new wastewater surveillance locations and modalities, and our results provide preliminary evidence for including certain methods when testing mNGS-based WBS. It is also critical to understand how methodological choices in a WBS mNGS workflow, including sample collection, extraction of nucleic acids, sequencing library preparation, sequencing platform and bioinformatic analysis of the resultant data affect operational LOD. There is currently no single gold-standard end-to-end workflow defined for mNGS, and variables throughout the workflow can introduce bias and impact results. ^61,62,63,64^ As mNGS approaches are more broadly deployed, there is a continued need to understand how workflow variables affect expected outcomes and performance in order to compare and contrast results from similar use cases.

Collectively, this study provides an example of how modeling and experimental approaches can be used to characterize the operational LOD for mNGS in wastewater samples. We provide insights into how the target concentration, background composition, bioinformatic algorithms and sequencing parameters affect the results of mNGS-based target detection. Subsequent work could include viral and Gram-positive bacteria spike experiments to further investigate the effects of process recovery and background noise. Other sample processing variations, such as depletion or enrichment, could be explored for potential operational LOD benefits or biases. Additional experiments in collaboration with other laboratories could be performed to assess sample-to-sample and lab-to-lab variability and gain more confidence in results. As WBS moves towards untargeted approaches like mNGS, the characterization of performance metrics using the process outlined here will be critical to gain decision maker confidence and enable actionable decision- making.

## Data Availability

All data produced in the present study are available upon reasonable request to the authors

## ACKNOWLEDGMENTS

We thank Stephen Francesconi (Defense Threat Reduction Agency) for helpful discussion and for sponsoring this work; Emily Kurdzo, Johanna Bobrow, Charlie Sinkler, Dave Walsh, Diane Jamrog and Henry Lau (MIT Lincoln Laboratory) for early thoughts on sample collection and processing; Jessie Hendricks (MIT Lincoln Laboratory) for advice on statistical analysis and permutation testing; David Wong and engineering team (Hanscom Air Force Base) for their help with sewer system maps and access to wastewater sampling sites; Nicholas Sevarino and Allison Heil (Weston and Sampson Engineers) for helping with the pilot dye test; Michael McLaren and William Bradshaw (SecureBio) for helpful discussion and advice on seeking permissions for spike experiments; Fred Sonnenwald and Ian Guymer (University of Sheffield) for helpful discussion on computational modeling.

## Contributions

AX Data Curation, Formal Analysis, Investigation, Software, Visualization, Writing KB Data Curation, Investigation, Writing

DC Data Curation, Investigation, Writing

TV Conceptualization, Investigation, Visualization, Writing

JS Data Curation, Formal Analysis, Investigation, Software, Visualization AM Data Curation, Formal Analysis, Software, Visualization, Writing

JL Conceptualization, Investigation, Visualization, Writing

DISTRIBUTION STATEMENT A. Approved for public release. Distribution is unlimited.

This material is based upon work supported by the Under Secretary of War for Research and Engineering under Air Force Contract No. FA8702-15-D-0001 or FA8702-25-D-B002. Any opinions, findings, conclusions or recommendations expressed in this material are those of the author(s) and do not necessarily reflect the views of the Under Secretary of War for Research and Engineering.© 2026 Massachusetts Institute of Technology.

Delivered to the U.S. Government with Unlimited Rights, as defined in DFARS Part 252.227-7013 or 7014 (Feb 2014). Notwithstanding any copyright notice, U.S. Government rights in this work are defined by DFARS 252.227-7013 or DFARS 252.227-7014 as detailed above. Use of this work other than as specifically authorized by the U.S. Government may violate any copyrights that exist in this work.

