## Supplemental Information for "Metagenomic Sequencing for Wastewater-Based Surveillance: Modeling and Experimental Approaches for Determining Limit of Detection"

DISTRIBUTION STATEMENT A. Approved for public release. Distribution is unlimited.

This material is based upon work supported by the Under Secretary of War for Research and Engineering under Air Force Contract No. FA8702-15-D-0001 or FA8702-25-D-B002. Any opinions, findings, conclusions or recommendations expressed in this material are those of the author(s) and do not necessarily reflect the views of the Under Secretary of War for Research and Engineering.© 2026 Massachusetts Institute of Technology.

Delivered to the U.S. Government with Unlimited Rights, as defined in DFARS Part 252.227-7013 or 7014 (Feb 2014). Notwithstanding any copyright notice, U.S. Government rights in this work are defined by DFARS 252.227-7013 or DFARS 252.227-7014 as detailed above. Use of this work other than as specifically authorized by the U.S. Government may violate any copyrights that exist in this work.

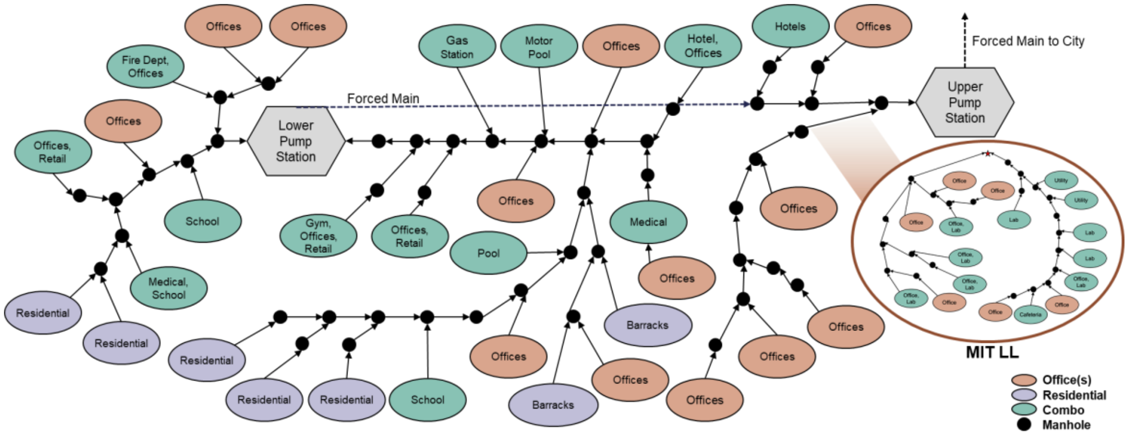


**Figure S1: Simplified routing network model of HAFB**. Locations draining into lower and upper pump stations are gravity fed. A forced main transports material from lower to upper pump wet wells and from there to a municipal line.

**Table S1: Water usage estimated by establishment type[1].**

| Establishment | Unit | Gallons per day |
|---|---|---|
| Residential dwelling | per bedroom | 110 |
| Hotel | per bedroom | 110 |
| Functional hall | per seat | 15 |
| Office building | per 1000 sq ft | 75 |
| Swimming pool | per person | 10 |
| Gymnasium | per participant | 25 |
| Hospital | per bed | 200 |

**Table S2: Primer and probe sequences for *Atlantibacter subterraneus* qPCR**

| **Target gene** | **Forward primer (5’->3’)** | **Reverse primer (5’->3’)** | **Probe (5’->3’)** | **Reference Fasta** |
|---|---|---|---|---|
| DedA | AACGCGCGAGAATAATGG | CTGTTCAGTAACCCGAACTC | 5’-/56-FAM/TTCCGCCGCAGCTATCTGGATAAA/3BHQ_1/-3’ | https://www.bv-brc.org/view/FASTA/dna/?in(feature_id,(PATRIC.255519.11.JAPTZM010000003.CDS.474670.475329.fwd |

**Table S3: Summary of estimated performance of three different sampling architectures.** Last column suggests the amount of concentration needed to reach theoretical mass inputs for the low shed case. EMI: estimated minimum input. *Assumption: sample volume = 300 mL, **High shed rate = 1×109 gc/g-feces, low shed rate = 1×107.53 gc/g-feces, †Branch architecture does not provide full coverage of source locations (based on physical parameters)

| **Architecture** | **Sample Number** | **Intercept Window** (qPCR, high shed**, 90th percentile) | **Peak Concentration** (High shed**) | **Scenarios Detectable by PCR (%)** (High shed**) | **Scenarios Detectable by Illumina Sequencing* (%)** (High shed**, Realistic EMI) | **Concentration Factor Required for Sequencing** (Theoretical EMI, Low shed**) | |
|---|---|---|---|---|---|---|---|
| | | | | | | **Illumina** | **MinION** |
| Base | 2 | 106 min | $1\times10^{5}$ gc/mL | 100 | 0 | $> 10^{3}$ | $> 10^{6}$ |
| Branch | 10 | ~ 5 min | $5\times10^{6}$ gc/mL | 78† | 0 | > 10 | $> 10^{4}$ |
| Building | 71 | ~ 2 min | $2\times10^{9}$ gc/mL | 100 | 57 | ~2 | $> 10^{3}$ |

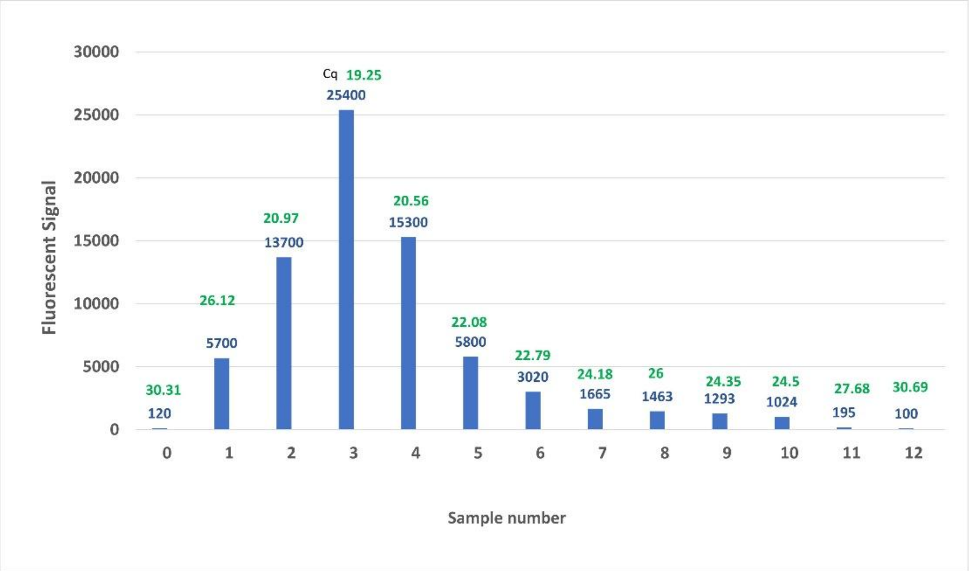


**Figure S2: Dye/Bacteria spike fluorescent intensity over time with corresponding Cq value.** After the system spike of Asub and initial 20 min transit time through the wastewater system, samples were taken at one min intervals. An aliquot of each sample was then analyzed by qPCR and fluorescent intensity. The Cq values (green) were from the Asub qPCR reaction for each sample. The fluorescent intensity values (blue) were readings from a fluorescent spectrometer (Horiba Duetta). Cq: quantification cycle

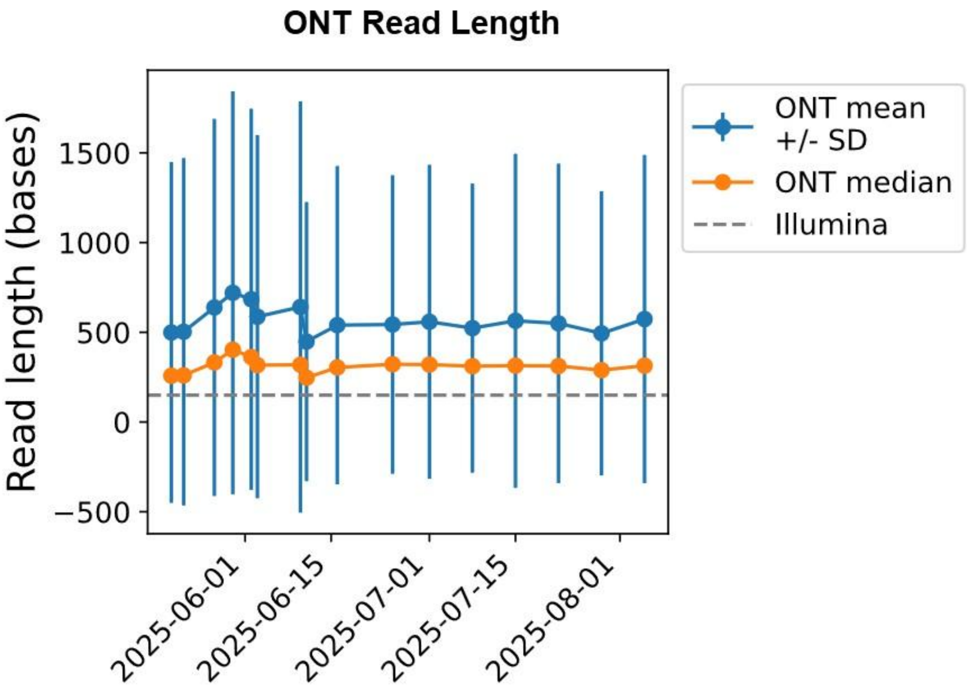


**Figure S3: Read length for Illumina and ONT after nucleic acid extraction and processing for selected wastewater samples.** Mean read length of ONT is shown in blue, with error bars corresponding to one standard deviation. Median read length of ONT is shown in orange. The 150 bp Illumina read length is shown in the gray dashed line.

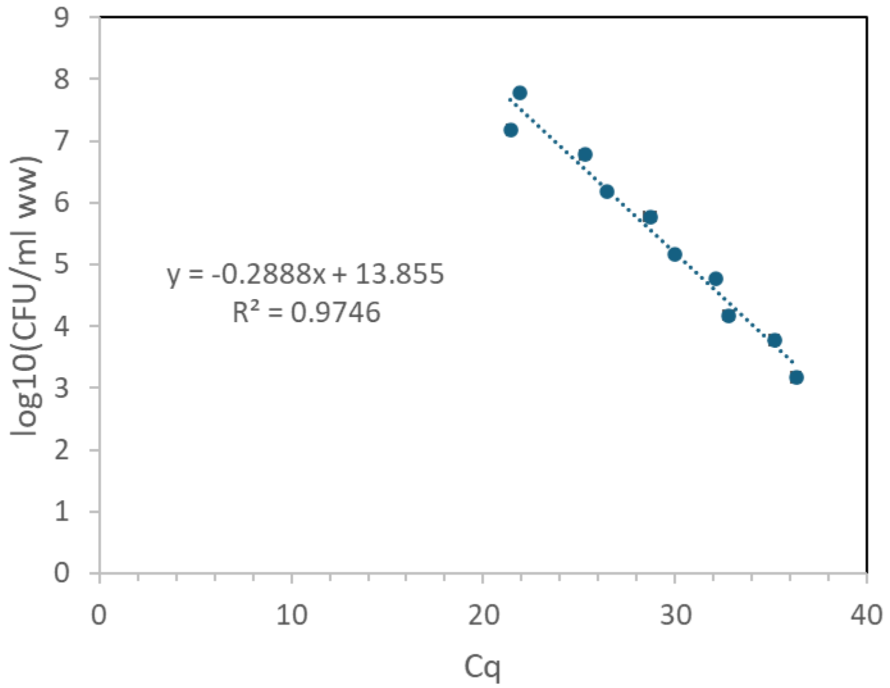


**Figure S4: Standard curve for interpolating concentration of system spike samples.** Asub whole cells were spiked into wastewater in the laboratory setting and taken through the extraction and qPCR process. A linear regression was used to build a standard curve equation to interpolate the concentration of system spike samples collected later. Samples with qPCR Cqs above the validated assay LOD Cq of 37.26 were excluded from the linear regression. Horizontal error bars represent the standard deviation of the Cq values. The linear regression equation log10(CFU/ml ww) = -0.2888 × Cq + 13.855 was used to estimate the CFU/mL ww of the system spike samples. CFU: colony forming units. WW: wastewater. Cq: quantification cycle.

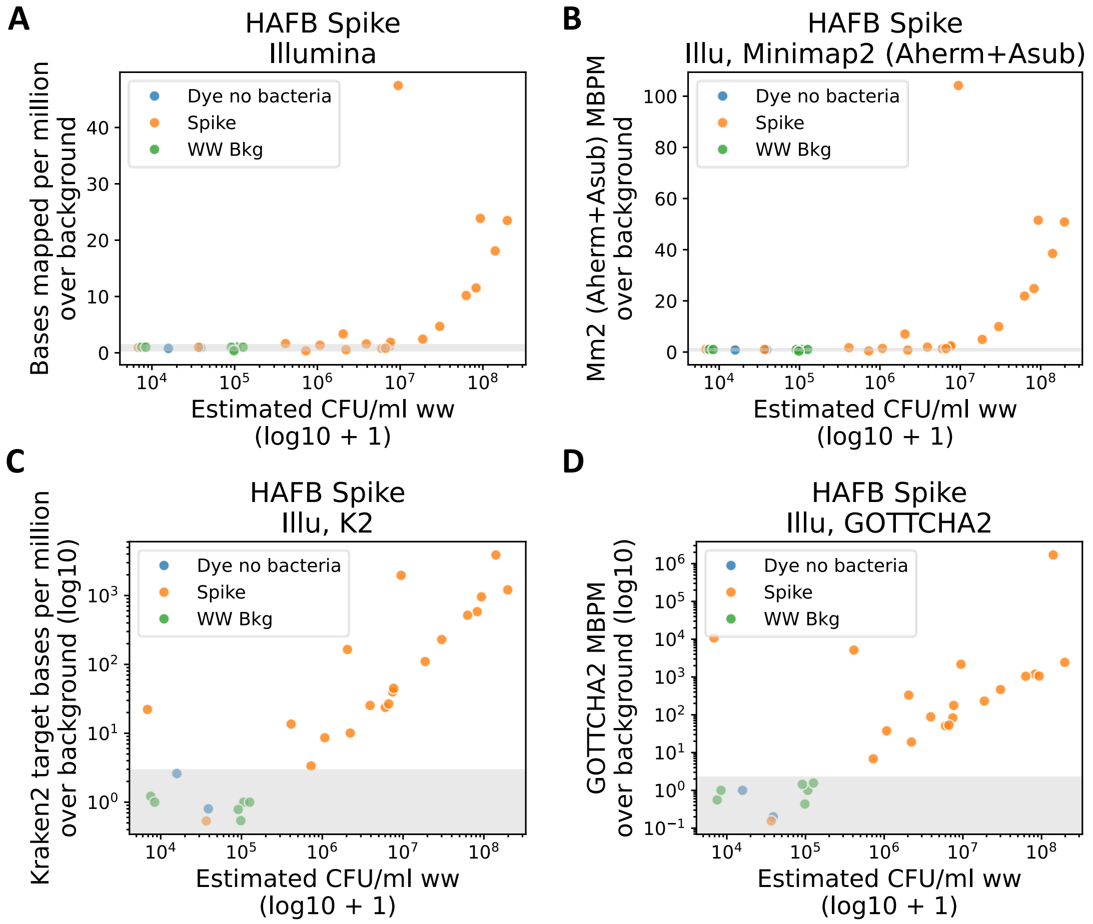


**Figure S5: Detection signals for Illumina data using different bioinformatic methods.** System spike data was analyzed with (A) minimap2 to the Asub reference genome, (B) minimap2 to a reference that included both Asub and *A. hermanii*, (C) Kraken2 with a broad database, and (D) GOTTCHA2 with a broad database. Gray region represents the average MBPM over background for non-spiked wastewater samples +/- 3 standard deviations. X-axis shows the estimated CFU/mL wastewater based on the qPCR calibration curve, with a pseudocount of 1 added to enable visualization of 0 on a log10 scale. CFU: colony forming units. WW bkg: wastewater background before target was spiked. Dye no bacteria: wastewater background with fluorescent dye only, without target bacteria. Spike: samples collected after target bacteria were spiked in the sewer system. MBPM: mapped bases per million, number of bases mapped to the target organism normalized by the number of bases after sequencing read quality control and multiplied by a factor of $10^6$. Asub: *Atlantibacter subterraneus*. Aherm: *Atlantibacter hermannii.*

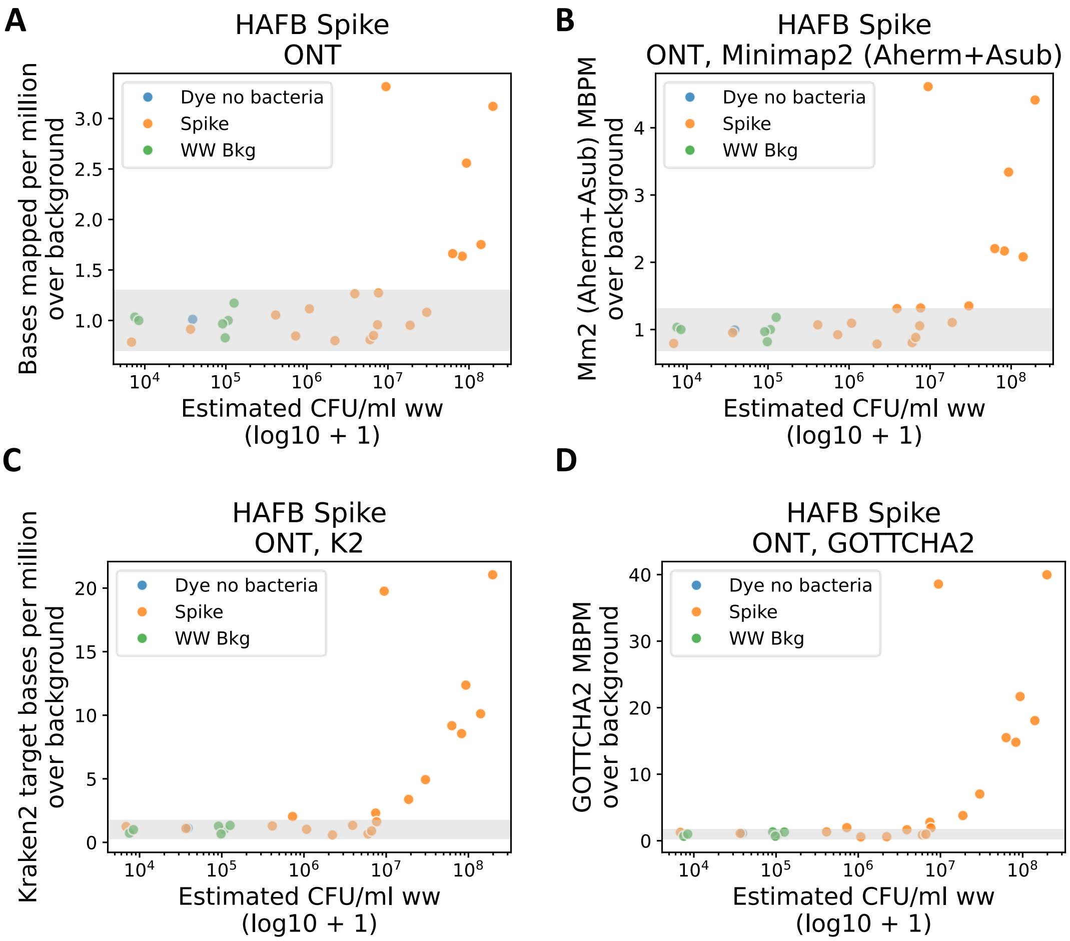


**Figure S6: Detection signals for ONT data using different bioinformatic methods.** System spike data was analyzed with (A) minimap2 to the Asub reference genome, (B) minimap2 to a reference that included both Asub and *A. hermanii*, (C) Kraken2 with a broad database, and (D) GOTTCHA2 with a broad database. Gray region represents the average MBPM over background for non-spiked wastewater samples +/- 3 standard deviations. X-axis shows the estimated CFU/mL wastewater based on the qPCR calibration curve, with a pseudocount of 1 added to enable visualization of 0 on a log10 scale. CFU: colony forming units. WW bkg: wastewater background before target was spiked. Dye no bacteria: wastewater background with fluorescent dye only, without target bacteria. Spike: samples collected after target bacteria were spiked in the sewer system. MBPM: mapped bases per million, number of bases mapped to the target organism normalized by the number of bases after sequencing read quality control and multiplied by a factor of $10^6$. Asub: *Atlantibacter subterraneus*. Aherm: *Atlantibacter hermannii.*
